# Geospatial Analysis to Determine Optimal Distribution of Mobile Stroke Units

**DOI:** 10.1101/2025.03.04.25323392

**Authors:** Ryan Ramphul, James C Grotta

## Abstract

**Introduction:** Mobile Stroke Units (MSUs) provide faster stroke treatment with improved outcomes but are expensive and their urban and rural deployment differs. Geospatial analysis may be useful for planning optimal MSU distribution.

**Methods:** We geo-coded Texas state-designated level-I or II stroke centers that did not overlap catchment areas and mapped 30, 60, 120 and 180-minute drive time buffers around each center, superimposing them on the distribution of stroke patients in the state including estimates of rural, vulnerable and minority populations within each buffer. We assumed that a MSU deployed from these “MSU centers” could rendezvous with EMS units halfway between a rural stroke location and the destination stroke center. For each buffer, we compared the number of patients potentially served by the MSU to a “base case” estimate of EMS transport represented by a 30-minute drive time buffer surrounding all non-overlapping level-I, II, III, or IV stroke centers.

**Results:** We identified 11 level-I and 3 level-II potential MSU stroke centers. A 180-minute buffer around each of these (MSU-EMS rendezvous 90 minutes from the stroke center) resulted in 741,852 stroke patients potentially receiving thrombolysis within 3 hours of stroke onset representing 99.1% adult stroke patients in the state; a net increase of 105,522 (16.6%) patients compared to “base case” and a 279% increase in patients from rural areas. A 120-minute buffer increased total and rural treatments by 12.3% and 232%. A 60-minute buffer resulted in no net increase in treated patients, though 600,101 more would receive faster care by MSUs.

**Conclusion:** When distributed using geospatial analysis, MSUs can provide faster acute stroke treatment and potentially better outcomes to virtually the entire state of Texas with a particular increase in rural populations that are not currently reached by EMS. Our findings might be useful to health care planners in any state.

## BACKGROUND

Management of acute stroke patients in Mobile Stroke Units (MSUs) compared to standard ambulance transport by emergency medical services (EMS) results in more frequent and faster treatment with intravenous thrombolytics (IVT) and better outcome as measured by disability 90 days after the stroke occurs. This benefit has been demonstrated in two large controlled clinical trials^1, 2^ and confirmed in a “real world” analysis of Get with The Guideline (GWTG) hospitals outside of a strict clinical trial protocol framework^3^. However, translating these data to widespread implementation of MSUs has several impediments, most importantly MSU cost and lack of reimbursement, and uncertainty about optimal MSU placement^4,5^. MSU placement in turn must consider population density and stroke prevalence, and the 3-4.5-hour IVT treatment time window that will constrain how far MSUs can travel to treat a patient. Geospatial analysis may be an ideal tool to address these issues and help determine where MSUs might be optimally implemented^6^.

To begin to address these implementation issues, one of the most important variables to consider is the number of patients that can be treated on MSUs. Cost benefit analyses demonstrate that the high fixed costs of operating a MSU are offset to a greater extent by the more patients that they treat^7^. MSUs in the U.S. treat on average approximately 50 patients per year with IVT^8^ which computes to first year post-stroke costs per Quality Adjusted Life Year (QALY) gained of $195,000 in patients with no pre-existing disability^7^. On the other hand, treatment of 150 patients per year, which is the case in the busiest MSUs, results in a cost per QALY of $61,000/yr.

The number of patients treated on MSUs will be influenced by their relative urban vs rural catchment area. MSUs included in the aforementioned MSU efficacy trials, GWTG study, and cost-effectiveness analyses operated primarily in urban communities with catchment areas of 10 miles or less radius from the MSU base station. In this scenario, the MSU, which arrives on scene at roughly the same time as the EMS ambulance, will evaluate and treat the patient on scene within 25 minutes of arrival^1^. If the patient were instead transported by EMS, the usual EMS transport time to the ED where the patient would be treated is also about 25 minutes. Therefore, the time saved by the MSU (30-45 minutes in most studies) is equal to the door to needle time in the Emergency Department (ED).

While the majority of stroke patients reside in urban areas, a considerable number, particularly those with the highest risk of stroke, reside in more rural areas^9^. The impact of MSU management in rural areas has been much less studied but would involve the “rendezvous” concept. With rendezvous, the closest EMS ambulance picks up the patient at a distant location and begins transport to the destination ED, while the MSU is dispatched from near the destination ED. The EMS ambulance and MSU rendezvous somewhere between where the patient is picked up and the destination ED, and the patient treated on the MSU at the rendezvous point. Rendezvous has been used successfully in some urban MSUs to extend their catchment area into suburban regions^10^. In a rural area, if the time of transport from the rendezvous point to the ED were greater than 25 minutes (e.g. the time it would take for the MSU to treat the patient), then the time savings by MSU management would be even greater than in urban areas. Thus, the use of rendezvous may result in a greater impact on outcome per patient in rural compared to urban areas. This has been demonstrated by the University of Florida MSU which services semi-urban Gainesville and 6 surrounding rural counties. In patients from the rural counties, the average dispatch to needle time for MSU management via rendezvous was 41.3 minutes compared to 97.8 minutes if patients had to travel by EMS to the stroke center, a net saving of 56.5 minutes^11^.

Geospatial analysis is increasingly used to analyze distribution issues such as optimal MSU placement that must take into consideration the location of stroke centers, stroke prevalence, population density, and time and distances travelled. In this study, we aimed to carry out a Texas statewide geospatial analysis to understand how to provide faster more effective MSU-based treatment to the largest number of Texas stroke patients if MSUs were strategically placed at level I or II state stroke centers, modeling various rendezvous time/distances to access patients. We hypothesized that MSUs via rendezvous could substantially increase the number of stroke patients receiving optimized treatment, particularly in rural regions of the state.

## METHODS

Texas established stroke designation rules in 2005 to create a state-wide emergency stroke treatment system, ensuring individuals showing acute stroke symptoms are transported to appropriately equipped facilities. These designations, updated in 2022 and similar to designations by national credentialing organizations, classify hospitals into four levels based on their capabilities: Level I (Comprehensive Stroke Centers), Level II (Advanced Stroke Centers, which can perform thrombectomy but are not comprehensive), Level III (Primary Stroke Centers), and Level IV (Acute Stroke Ready Centers).

Using ArcGIS Pro Version 3.3.2 {https://www.esri.com/en-us/arcgis/products} we geocoded all level I and II stroke facilities. Data on stroke facilities came from the Texas Department of State Health Services. For strategic placement of MSUs, we included only level I and II centers because it is unlikely that MSU operations would be supported by a level III or IV center and wanted to demonstrate the impact if all non-overlapping Level I and II centers would have MSU programs. Since MSU programs should optimally be coordinated among all the stroke centers in a given community or region, we modeled only 1 MSU if the level I or II stroke centers were located within 30 minutes of each other. Next, we created a series of drive time buffers around each of the “MSU centers” included in our analysis, using ArcGIS Pro. A drive time buffer is a polygon shape of how far one can drive in any direction from each input stroke facility, using the underlying road network, in the specified period.

We created a 30-minute drive time buffer as the baseline representing the usual transport time for a standard EMS ambulance, and then one-hour, two-hour, and three-hour drive time buffers to estimate the time and distance travelled if the MSU rendezvoused with a EMS unit inbound from a distant scene. Assuming MSU rendezvous with EMS halfway between the site of the stroke and the destination level I or II center, the one, two and three-hour buffers would correlate with MSU drive times of 30, 60, or 90 minutes. Although guideline-approved to 4.5 hours, optimal administration of IV thrombolysis should occur within 3 hours of onset. Assuming 1.5 hours for stroke recognition, EMS alerting and arrival, and evaluation and treatment on the MSU upon rendezvous, we assumed that a MSU drive time of up to 90 minutes from MSU base to rendezvous (e.g. the 180-minute buffer) might still allow MSU-based treatment within the 3-hour time window.

For comparing MSU stroke coverage to current EMS coverage (i.e. “base case”), we created 30-minute drive time buffers surrounding all non-overlapping level I, II, III, or IV state stroke centers (n= 190). This assumed 1.5 hours for stroke recognition, EMS alerting arrival and evaluation on scene, 30 minutes EMS transport time, and 1 hour door to needle time in the stroke center ED.

### Estimating adult populations within different drive times from stroke centers

We estimated the number of adults (18+) within each drive-time buffer around stroke centers using data from the CDC PLACES 2024 Release, which provides model-based estimates of chronic disease indicators at the census tract level. The CDC PLACES dataset is a robust and nationally consistent resource developed by the CDC in collaboration with the Robert Wood Johnson Foundation and the CDC Foundation. It uses advanced small area estimation methods to provide health indicator estimates for all U.S. census tracts, allowing for detailed, localized public health assessments even in areas lacking comprehensive survey data. Specifically, we summed the estimated number of adults in each tract whose centroid fell within each respective drive-time buffer (e.g., 30 minutes). Drive-time zones were generated using GIS-based network analysis, which accounts for real-world road networks and travel speeds to more accurately reflect access to care.

### Estimating stroke prevalence within different drive times from stroke centers

To estimate stroke prevalence within each buffer, we used tract-level stroke prevalence estimates from the CDC PLACES dataset (Figure 1). Rather than applying a uniform statewide stroke prevalence, we calculated a population-weighted average stroke prevalence for each drive-time area. This was done by multiplying each tract’s stroke prevalence rate by its estimated adult population, summing the results across all included tracts, and dividing by the total adult population within the buffer. This method allows for a more precise, geographically sensitive measure of stroke burden relative to proximity to stroke care.

**Figure 1:**
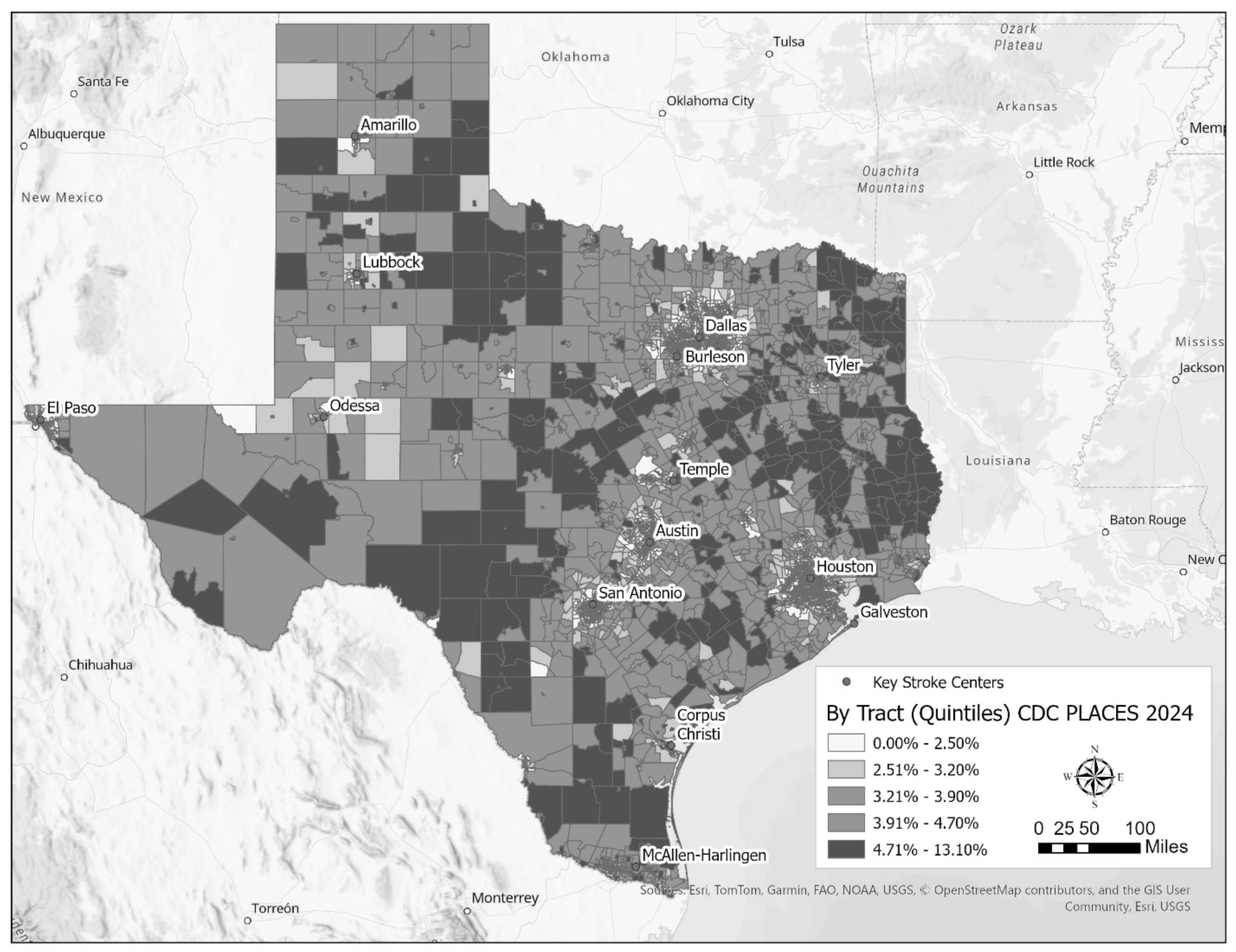
Stroke Prevalence by Census Tract CDC PLACES 2024.

### Estimating rural populations within different drive times from stroke centers

Next, we utilized Rural-Urban Commuting Area (RUCA) codes to estimate rural populations within different drive times from stroke centers. Rural-Urban Commuting Area (RUCA) codes, developed by the U.S. Department of Agriculture, classify geographic areas, in this case census tracts, based on urbanization, population density, and commuting patterns. These codes range from 1 to 10, with lower scores indicating more urban areas and higher scores representing more rural regions. To simplify analyses, researchers frequently group RUCA codes into broader categories to differentiate these settings more effectively and facilitate studies on health disparities, resource allocation, and access to services. As such we grouped RUCA Codes into three categories: urban, rural, and micropolitan (or small-town), in the following manner, based on guidance from the USDA:

- **Urban**: RUCA codes 1 through 3, represent areas with high population density and strong commuting ties to urban centers.
- **Micropolitan**: RUCA codes 4 through 6, covers smaller towns or regions with moderate commuting ties to urban areas.
- **Rural**: RUCA codes 7 through 10, indicate areas with low population density and weak commuting ties to urban centers.

This grouping helps streamline analyses and comparisons while maintaining meaningful distinctions between types of geographic areas. To ensure accuracy, we removed census tracts containing airports, which are often assigned a RUCA score of 10 due to low residential population, but do not represent truly rural areas. To estimate rural populations reachable from different drive time buffers from stroke centers, we calculated the adult populations in rural census tracts (RUCA codes of 7-10) within each buffer. Because of their distance from stroke centers, we assumed that rural patients would be unlikely to receive IVT within 3 hours of stroke onset by current EMS transport.

### Estimating socially vulnerable populations within different drive times from stroke centers

To estimate socially vulnerable populations within varying drive times from stroke centers, we utilized the CDC’s 2020 Social Vulnerability Index (SVI). The CDC’s Social Vulnerability Index (SVI) is a tool that uses U.S. Census data to measure the relative social vulnerability of communities based on 19 variables, such as poverty, access to transportation, and housing conditions. These variables are grouped into four themes—socioeconomic status, household composition, minority status and language, and housing/transportation—and combined to produce a percentile ranking for each census tract, with higher scores indicating greater vulnerability. The SVI helps identify areas that may require additional support during public health emergencies or resource allocation efforts.

To estimate especially vulnerable populations, we isolated census tracts with SVI percentile scores of 0.75 or higher, representing the most vulnerable populations. We aggregated the adult populations (18+) in these high-SVI census tracts whose centroids fell within the 30-minute, 1-hour, 2-hour, and 3-hour drive time buffers around stroke centers. This method highlights areas where socially vulnerable populations may face significant barriers to accessing timely stroke care, informing strategies for improved healthcare equity.

### Estimating minority populations within different drive times from stroke centers

To estimate minority populations within different drive times from stroke centers, we used census tract-level estimates of the percentage of non-White populations provided in the CDC’s 2020 Social Vulnerability Index (SVI) data package. We aggregated the adult populations (18+) in census tracts based on the proportion of non-White residents and whose centroids fell within the 30-minute, 1-hour, 2-hour, and 3-hour drive time buffers. This approach allows for a detailed assessment of disparities in stroke care accessibility among minority populations, offering crucial insights into areas that may require targeted interventions to reduce health inequities.

## RESULTS

Using tract-based estimates from the CDC PLACES dataset, we estimate that there are approximately 748,835 adult stroke patients in Texas of whom an estimated 49,090 reside in rural areas and therefore currently unlikely to receive acute stroke treatment with IVT within 3 hours of stroke onset. Additionally, data from the CDC PLACES project indicates considerable stroke geographic heterogeneity, with high prevalence in rural regions particularly in East Texas and along the Mexican border (Figure 1). Using the criteria described in the Methods, we identified 11 potential MSU locations with level I facilities, and 3 with level II (Amarillo, Odessa, and Burleson) (Figure 2). Superimposing figure 2 on figure 1 provides a visual representation of the proportion of stroke patients that might be served by MSUs using the rendezvous approach. The data are presented in Table 1.

**Figure 2:**
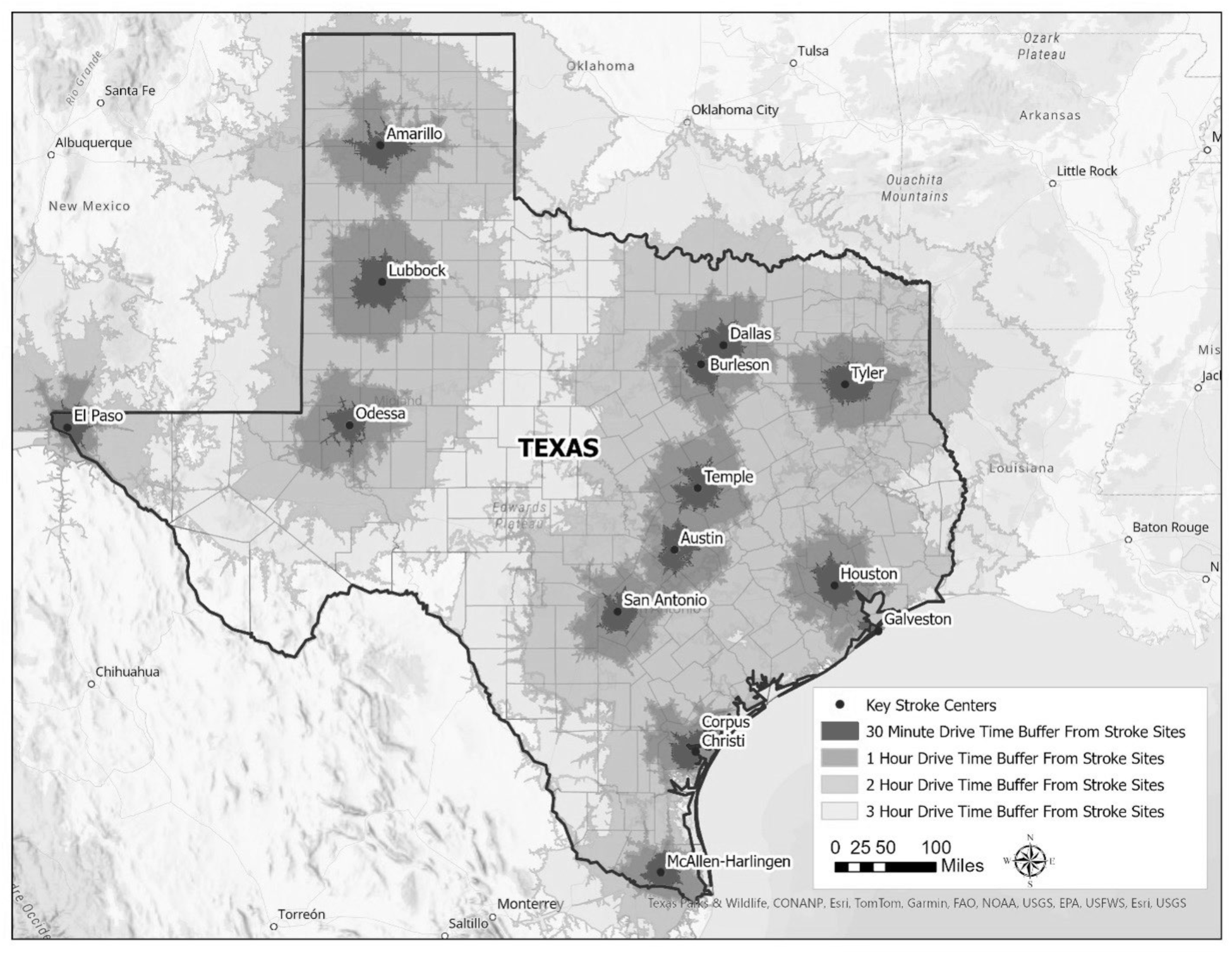
Drive Time Buffers Around Potential MSU Equipped Level I or II Stroke Facilities.

**Table 1:** Modeling of Additional Stroke Patients That Could Be Served.

| **Drive Time Radius Around Stroke Centers in Texas** | **0.5 Hour Baseline from ALL Stroke Centers (Population-Weighted Mean Stroke Rate 3.3%)** |  | **1 Hour From 14 Key Stroke Centers (Population-Weighted Mean Stroke Rate 3.3%)** |  |  | **2 Hour From 14 Key Stroke Centers (Population-Weighted Mean Stroke Rate 3.4%)** |  |  | **3 Hour From 14 Key Stroke Centers (Population-Weighted Mean Stroke Rate 3.4%)** |  |  |
| --- | --- | --- | --- | --- | --- | --- | --- | --- | --- | --- | --- |
|  | **Est.** | **Est. With Stroke** | **Est.** | **Est. With Stroke** | **Increase From Baseline (%)** | **Est.** | **Est. With Stroke** | **Increase From Baseline (%)** | **Est.** | **Est. With Stroke** | **Increase From Baseline (%)** |
| Adult Pop > 18 | 19,282,722 | 636,330 | 18,184,885 | 600,101 | N/A | 21,024,426 | 714,830 | 78,500 (12.3%) | 21,819,178 | 741,852 | 105,522 (16.6%) |
| Adult Pop in Census Tracts Designated as "Rural" | 288,767 | 9,529 | 200,521 | 6,617 | N/A | 929,686 | 31,609 | 22,080 (231.7%) | 1,060,958 | 36,073 | 26,544 (278.6%) |
| Adult Pop in Census Tracts in the Highest Percentile Score of Social Vulnerability | 4,514,318 | 148,972 | 4,262,327 | 140,656 | N/A | 4,845,957 | 164,763 | 15,791 (10.6%) | 5,093,848 | 173,191 | 24,219 (16.3%) |
| Adult Minority Pop | 11,408,218 | 376,471 | 10,906,383 | 359,911 | N/A | 12,051,531 | 409,752 | 33,281 (8.8%) | 12,495,090 | 424,833 | 48,362 (12.8%) |

A one-hour drive time radius around each of the 14 MSU centers in our sample (MSU-EMS rendezvous 30 minutes from the stroke center) resulted in no net increase in the estimated number of stroke patients that could be treated on MSUs compared to the number treated following transport by EMS to the 190 level I-IV stroke centers around the state within a 30-minute drive time. However, the 636,330 patients treated on MSUs would be treated faster and consequently have a greater chance of good outcome compared to if they were transported by EMS^1,2^

By increasing the MSU service area to a two-hour drive time radius around each of the 14 MSU centers in our sample (MSU-EMS rendezvous 1 hour from the stroke center), an estimated 714,830 patients could receive faster more optimized treatment on a MSU, including a net increase of 78,500 or 12.3% more stroke patients treated with IVT than with existing EMS coverage. About 40% of this increase (31,609 patients) would be from rural areas, which is a 231.7% increase compared to existing EMS coverage. Roughly equal numbers though proportionately smaller increases would occur in socially vulnerable and minority patients treated.

Finally, by increasing the MSU service area to a three-hour drive time radius around each of the 14 MSU stroke centers in our sample (MSU-EMS rendezvous 90 minutes from the stroke center), an estimated 741,852 patients representing 99.1% of all estimated adult patients suffering from stroke in the state could receive faster more optimized treatment on a MSU, including a net increase of 105,522 or 16.6% more stroke patients treated with IVT than with existing EMS coverage. Further proportionate increases were seen in rural, socially vulnerable and minority populations.

## DISCUSSION

Our data demonstrate that strategic location of MSU programs linked to 14 level I or level II stroke centers in Texas could substantially increase the number of stroke patients receiving faster more optimized MSU-based IVT treatment within 3 hours of stroke onset, reaching 99.1% of stroke patients in the state. This would have a particularly striking impact on increasing coverage of rural areas where, as seen in Figure 1, stroke prevalence is highest and where patients must travel great distances to reach a major stroke center. Currently rural patients with stroke are less likely to receive IVT or endovascular therapy and have higher in-hospital mortality than their urban counterparts^12^. One problem with new technological advances is that once implemented they are often not as available to remote, socially vulnerable and minority populations. Our data demonstrate that this would not be the case with geospatially guided MSU implementation.

Our findings, though based on modeling of Texas stroke centers and population density, might be useful to health care planners in any state. MSUs are a beneficial cost-effective innovation but are expensive and labor intensive to implement and operate. Communities must therefore decide if they can support a MSU, and health care planners must decide where they should be optimally located to deliver faster and more frequent treatment to patients who are located a substantial distance from the destination stroke center. In Texas, legislation has been introduced to provide funding for new MSU programs. Such support is essential until appropriate reimbursement for MSU services is provided by health care payers. Our geospatial analysis could be useful for the Texas legislature and for any state or health care planner to understand where MSU support might be best targeted.

MSUs must be licensed and staffed with sophisticated equipment such as CT scanners, and nursing and technological personnel familiar with stroke diagnosis and treatment. Therefore, all MSUs in current operation in the U.S. are operated by hospital systems in collaboration with local EMS. Level I stroke centers provide the full range of stroke treatments and are the current “home” for all MSUs in the U.S. However, level II centers can provide all stroke treatments including thrombectomy so might be able to support a MSU program in an area where a level I center is not available. These considerations guided the selection of the stroke centers where MSU programs would be located in the state for our analysis.

In urban areas, level I stroke centers tend to cluster and overlap catchment areas. It would not be efficient for each level I center to operate a MSU. Instead, in all communities with MSUs in the U.S., the MSU is operated by one level I center, but collaborates with others. Since the average EMS transport time to level I centers in urban areas is approximately 20-30 minutes, we considered all level I and II stroke centers as a single center if they were within 30 minutes of one another. Even if more than one stroke center within that overlapping area were to operate a MSU, the distances traveled to rendezvous would be the same for both so this would not result in a net increase in patients treated outside the 30 minute drive time buffer. In population-dense urban areas, there would be a need for more than one MSU whether managed by one level 1 center or several. If each of MSU centers in the state had at least two MSUs so that one would be available for rendezvous from the surrounding ex-urban areas while the others were available for the more urban area, we calculate that a total of approximately 34 MSUs could provide faster optimized MSU-based stroke treatment to the entire population of the state. This calculation assumes a total of 4 MSUs in the Houston and Dallas metro areas, 3 in San Antonio and Austin, and 2 in each of the other 10 MSU centers identified for this study.

As with any modeling exercise, we have made several assumptions, and the main limitation of our study is that it is based on these assumptions. While MSUs and EMS ambulances in urban areas usually have a roughly 10-mile radius catchment area, corresponding to an approximate 20-30-minute drive time, MSUs can cover a much larger area by using the rendezvous approach. With rendezvous, the MSU meets the EMS squad travelling with the patient from the distant site of the stroke, usually about halfway between the MSU base station and the site of the distant stroke, and treats the patient at the rendezvous site. For our study, we used the 30-minute drive time as the current “base” condition for both EMS and MSUs, and modeled MSU catchment areas based on drive times of 60, 120, and 180 minutes from the receiving stroke center, corresponding to MSU drive times of 30, 60, and 90 minutes from MSU base to rendezvous. Although guideline-approved to 4.5 hours from stroke onset, the benefit of IVT quickly decays with treatment beyond 1 hour^13^ and is FDA approved within 3 hours. Therefore, we used the 3-hour time window as the goal for IVT delivery for this study. Assuming 1 hour for stroke recognition and EMS arrival and evaluation on scene, and 30 minutes for treatment on the MSU upon rendezvous, an MSU drive time of over 90 minutes from MSU base to rendezvous (over 180 minutes total transport time) would probably put the patient outside the 3-hour time window. In more remote regions, certainly MSUs could travel longer distances but would not likely lead to treatment within 3 hours. If we changed our analysis to allow treatment within 4.5 hours from stroke onset, if anything this would increase the relative advantage of MSU over standard transport. The main reason patients are treated between 3-4.5 hours rather than within 3 hours is that they delay calling 911. This habit should occur with equal frequency whether the patient is transported by MSU or by standard ambulance. However, the 4.5-hours time window would enable the MSU to travel a longer distance to rendezvous and still be able to treat the patient. This advantage would not occur with standard ambulance operation where transport is confined to a more constrained distance from the local stroke centers. If a patient outside that catchment area delayed calling 911, they would have to be first triaged to a non-stroke center, evaluated there and then secondarily transported by ground or air to the stroke center, a process that would certainly take more than 90 additional minutes and therefore putting them outside the 4.5-hours time window. Interestingly, a recent analysis showed that stroke treatment of remote patients transported by helicopter ambulance is slower than ground transport^14^ so it is unlikely that air transport would be faster than EMS-MSU rendezvous except for exceptionally long distances. Our results correspond well with findings from Gainesville which is the only existing MSU program with published data on rural populations^11^.

We also had to make assumptions about how to compare projected MSU coverage to existing coverage by EMS without MSUs. All Texas state level I-IV stroke centers can treat stroke patients with IVT. Therefore, for our base case scenario, we used a 30-minute drive-time catchment area surrounding all 190 level I-IV stroke centers in the state. We used the 30-minute catchment area since a 20-30-minute drive time is average for most EMS units transporting stroke patients.

Regarding limitations in our geospatial analysis, population data were derived from CDC PLACES and other publicly available datasets, which are subject to sampling variability, especially in rural or low-density areas. These estimates may not precisely reflect real-time population dynamics, particularly in rapidly growing or declining communities. Second, our analysis assumes uniform road conditions and average travel times, not accounting for real-world variability such as traffic congestion, road closures, or geographic barriers (e.g., rivers or mountains), which may affect actual MSU travel times. Third, the modeling does not incorporate temporal factors such as time of day or day of week, which can influence EMS and MSU response and transport times. These factors could impact the precision of our coverage estimates and the generalizability of our findings to real-world MSU deployment scenarios.

In conclusion, using geospatial modeling to optimize their strategic location, MSUs can provide faster more effective stroke treatment to the vast majority of stroke patients in Texas, including rural, socially vulnerable and minority populations, including rural, socially vulnerable and minority populations. MSU rendezvous with EMS travelling from 2 hours away will increase the number of rural patients receiving treatment by 231%, and from 3 hours away could provide faster optimized MSU-based treatment for 99.1% of all stroke patients in the state.

## Data Availability

The data used in this study were obtained from publicly available sources, including the Texas Department of State Health Services, the 2020 U.S. Census American Community Survey (ACS) 5-year estimates, the CDC PLACES project, and the CDC Social Vulnerability Index (SVI). Geospatial analyses were conducted using ArcGIS Pro. Any additional data supporting the findings of this study are available from the corresponding author upon reasonable request.

